# Sample Size and Power Calculations Based on Endpoint Responsiveness: Applications to Trials of Disease-Modifying Therapies in Parkinson’s Disease

**DOI:** 10.64898/2026.09.29.26363876

**Authors:** Miriam Pedrera Gomez, Corrado Bernasconi, Claudia Mazzà, Shibeshih Belachew

## Abstract

In Parkinson’s disease (PD), early phase clinical trials of disease-modifying agents generally use the MDS-UPDRS Part III as the primary outcome measure of motor function, which typically requires a large sample and/or a long (12– 18 months) treatment duration, even if a substantial treatment effect is assumed. This is due to the rather slow rate of early PD progression relative to the variability of the MDS-UPDRS Part III assessments. Using placebo-arm data from four recent large Phase 2 trials of early-stage PD (SPARK, PASADENA, PADOVA and LUMA), the sample size needed to detect a range of assumed treatment effects on MDS-UPDRS Part III progression was estimated. The trial sample size estimates obtained for MDS-UPDRS Part III and for smartphone-based sensor-derived motor functional composite (MFC) endpoints were compared using an approach based on relative responsiveness to disease progression. The calculations were also implemented in an interactive, publicly accessible application. Powering a one-year trial using the MFC_1_ endpoint, derived from three active assessments (6-minute walk, U-turn and Hand Pronation-Supination), or the MFC_2_, which adds 60-Second Typing, was projected to reduce the required sample size by more than half relative to the MDS-UPDRS Part III, assuming equivalent proportional treatment effects.

## I introduction

The Movement Disorder Society Unified Parkinson’s Disease Rating Scale (MDS-UPDRS), a multi-domain clinician/patient-rated ordinal scale, is currently the most used clinical measure in Parkinson’s disease (PD) drug trials [1]. Nonetheless, the variability in motor and non-motor symptoms, as captured by this instrument, limits the sensitivity of trials to detect changes in disease severity and treatment effects. This is especially true in the early stages of PD, where the rate of change on the MDS-UPDRS scale is small relative to the intra-subject variability [2], [3]. This implies that treatment studies of disease modification require a very large sample size, or as a less practicable approach in drug development, a very long treatment duration.

The aim of this paper is to quantify this trial-design challenge, while investigating the potential of alternative candidate endpoints with higher longitudinal responsiveness. For this purpose, the total sample size required to detect a treatment effect on MDS-UPDRS Part III progression in a hypothetical randomized controlled trial was estimated using placebo arm data from four recent controlled trials in early PD.

This work also examines whether smartphone-derived motor functional composite (MFC) endpoints with higher longitudinal responsiveness could improve statistical power and reduce sample size requirements when compared to the MDS-UPDRS Part III. Two composite endpoints, MFC_1_ and MFC_2_, were developed and evaluated in LUMA, a placebo-controlled Phase 2b study of BIIB122/DNL151 [4], based on a prespecified statistical analysis plan including a validation in an independent held-out dataset. MFC_1_ combines measures of walking, turning, and hand movement (6-minute walk, U-turn and Hand Pronation-Supination); MFC_2_ adds a typing measure (60-Second Typing). There was only a marginal difference in responsiveness between those two composite endpoints [4]. The analyses focus primarily on MFC_1_, whose content reflects domains assessed by the MDS-UPDRS. A third smartphone-derived composite, the Simple Sum v1.2 Score [5], is included as an additional illustration of the approach.

## IImethods

### A. Sample Size Estimation for a Reference Endpoint

The example case for this analysis is a controlled clinical trial of a disease-modifying agent using a continuous clinical outcome measure, which, in a placebo arm, is expected to increase linearly over time reflecting the natural rate of disease progression. The disease-modifying intervention in the trial is expected to slow down that rate of progression.

Accordingly, the treatment effect can be expressed as the percent slope reduction of the outcome measure in the active treatment arm compared to placebo.

Cohen’s *d* effect size for a given assumed slope reduction is then obtained using the standardized response mean (SRM)

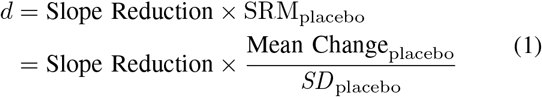

*SD* _placebo_ is used here instead of *SD* _pooled_ because data from the active treatment arm were not available for all cohorts. The underlying assumption is that the endpoint variability is independent of the mean, at least in the investigated range of treatment effects.

Two illustrative hypothetical scenarios are considered: Drug A, leading to a 25% slope reduction, and Drug B, leading to a 50% slope reduction. In addition, where a source reported active-arm values at the same time point, the observed treatment effect was computed as:

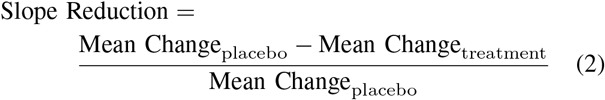

For the sample size, estimated with the `pwr.t.test` function of the `pwr` package (version 1.3-0) [6] in R (version 4.6.1), 80% statistical power is targeted for a two-sided *t*-test at the 5% significance level. Where an allocation ratio other than 1:1 is specified in the calculator, power is evaluated for the resulting unequal group sizes with `pwr.t2n.test`.

This reflects a between-arm comparative analysis of the mean change of the outcome at one year. Typically, in an actual future study, this endpoint would be evaluated by a longitudinal linear model. Sample size results are expressed as the number of evaluable patients, i.e. disregarding the number of patients who are randomized but not eligible for the analysis (typically rather small for the longitudinal analysis of the relevant study types).

### B. Responsiveness ratio to calculate an endpoint’s effect size

A candidate outcome measure may be considered as a potential replacement for a reference endpoint on the grounds of a higher longitudinal responsiveness. The responsiveness ratio, i.e. the ratio of the candidate SRM to the reference SRM, can be used to convert an assumed effect size for the reference endpoint to an effect size for the candidate endpoint. To avoid contamination by any treatment effect on the mean change from baseline, the responsiveness ratio is computed using data from the placebo arm.

In practice, for any given time point of interest, Cohen’s effect size for the reference endpoint is multiplied by this responsiveness ratio:

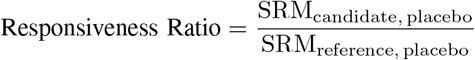

And

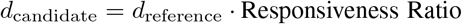

where *d* is defined as per (1) and the assumption is made that the two measures have a linear change from baseline and the same slope reduction, so that the between-arm difference (Δ) in the change from baseline for the candidate can be calculated as:

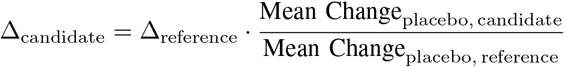

The MDS-UPDRS Part III serves as the reference endpoint for these analyses, with MFC_1_ as the principal candidate alternative and MFC_2_ included in the illustrative case study. Both composites were developed in the LUMA Phase 2b study [4].

MFC_1_ combines four sensor-derived measures from three smartphone-based assessments: the average wrist rotation amplitude across hands from the Hand Pronation-Supination assessment, the 95th percentile turning speed from the U-turn assessment, and the total distance walked and median walking cadence from the 6-minute walk. MFC_2_ includes these four measures plus the 5th-percentile letter interval from the 60-Second Typing assessment. Both composites are calculated by summing sample-based z-scores for their constituent measures.

### C. Trial Data Sources

Sample size considerations are developed here for future trials with a primary endpoint assessed at one year, a typical scenario for drugs evaluated for proof-of-concept of Parkinson’s disease modification in Phase 2. Published information was identified for the placebo arm of four controlled PD trials with slightly different patient populations (Supplementary Table S1):

- **SPARK** [7] (NCT03318523) was a Phase 2 study of cinpanemab in early PD patients who were untreated with symptomatic medication at study entry. Week 52 values were taken from the trial results disclosure, with the higher-dose arm (*N* = 51) used for the treatment effect. In that active treatment arm, the mean change was marginally larger than under placebo, so that the corresponding treatment effect estimate would be negative.
- **PASADENA** [8] (NCT03100149) was a Phase 2 study of prasinezumab in patients with early PD who had received no previous symptomatic treatment, except for monoamine oxidase B inhibitors. Placebo-arm and active-arm values at Week 52 were taken from the trial results disclosure, the treatment effect being that of the low-dose arm. A smartphone-derived Simple Sum v1.2 Score was also derived in the placebo arm and is discussed below.
- **PADOVA** [9] (NCT04777331) was a Phase 2b study of prasinezumab in early PD in which all participants were on stable symptomatic monotherapy (levodopa or a monoamine oxidase B inhibitor) at entry. Week 48 values were extracted from Figure 4 reported in Nikolcheva et al. [9] for the placebo arm (*N* = 272) and for the active arm (*N* = 273).
- **LUMA** [4] (NCT05348785) was a recent placebo-controlled Phase 2b trial of the LRRK2 inhibitor BIIB122/DNL151 in idiopathic early-stage PD. It is the cohort in which a candidate endpoint evaluated in this paper, the MFC_1_, was developed. Because MFC_1_ was reported concurrently with the MDS-UPDRS Part III in the same placebo arm, MDS-UPDRS Part III will serve as an anchor for the responsiveness ratio. The Week 48 SRMs in the placebo arm were taken directly from the published report. No data on the treatment effect observed in LUMA, on either the MDS-UPDRS Part III or the MFCs, are publicly available; only the communication that the trial was negative has been made.

From these sources, the mean and the variability of the change from baseline in the MDS-UPDRS Part III total score at approximately one year were extracted (Table I); for LUMA, only the placebo-arm SRM itself was available. If available, the treatment effect was also computed according to (2). For comparability, the reported values closest to one-year post-randomization were used (48–52 weeks: Week 52 for SPARK and PASADENA, Week 48 for PADOVA and LUMA). For PASADENA, the Week 52 Simple Sum v1.2 Score SRM was extracted from a linear model in the placebo arm reported in Taylor et al. [5]: the adjusted mean change (4.4) and the standard error (0.6) were read off the published figure, and the standard deviation was back-calculated from the standard error and sample size (*n* = 52). The standard error is the midpoint of two independent readings of the figure; how it was obtained, and the sensitivity of the results to it, are described in the Supplementary Materials.

## III. Results

### A. Sample Size Estimates

For each of the four trials, Fig. 1 displays the total sample size required to detect an assumed treatment effect (expressed as a slowing of the rate of MDS-UPDRS Part III worsening) in a placebo-controlled study. The same assumed treatment effects were also translated into the sample size required if the MFC_1_ endpoint were used in place of the MDS-UPDRS Part III.

**Fig. 1.**
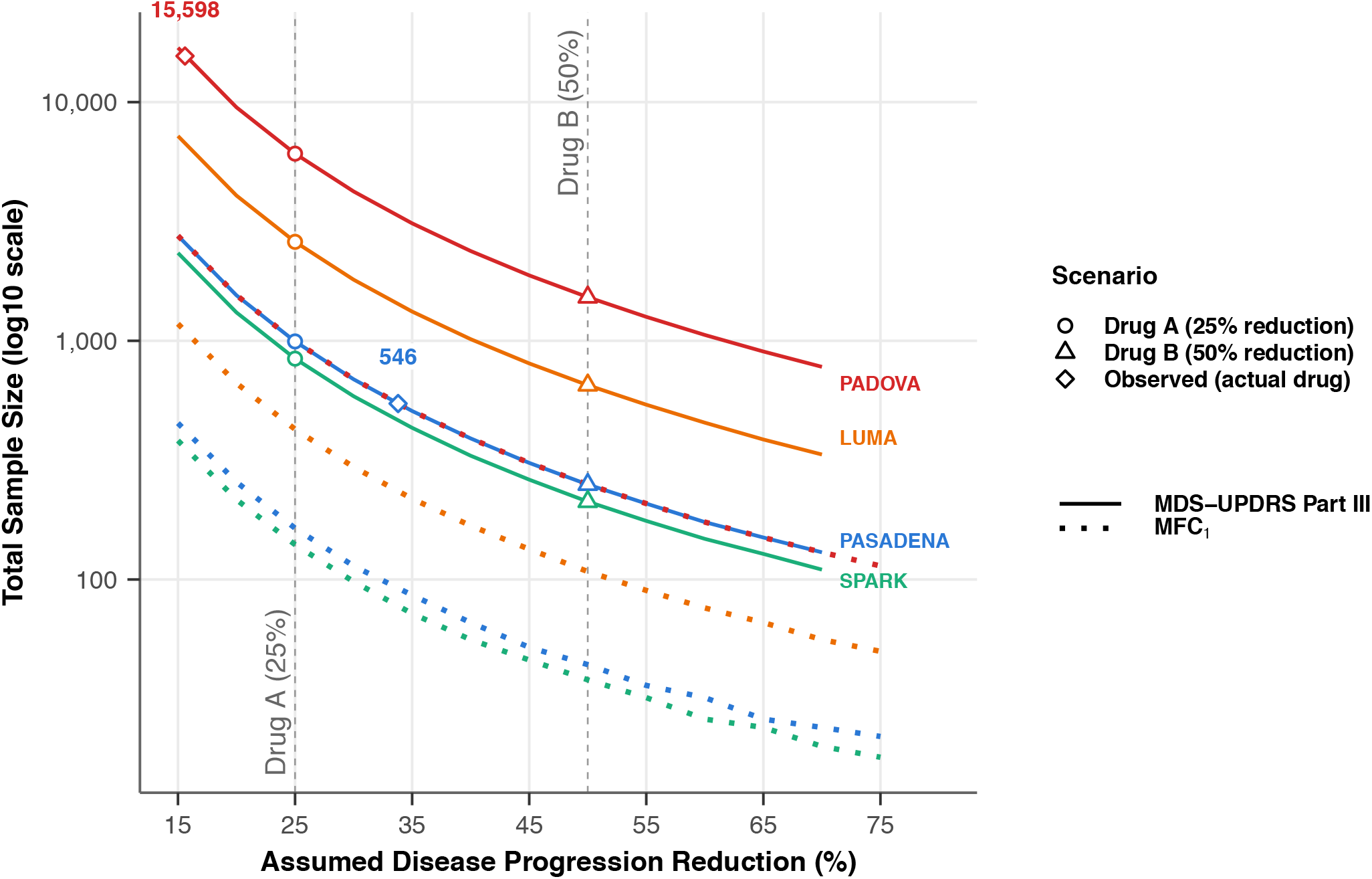
Total sample size (evaluable subjects) required to detect a range of assumed treatment effect-related reduction in slope of disease progression, by trial cohort, for the MDS-UPDRS Part III (reference endpoint, solid lines) and for the MFC_1_ candidate alternative endpoint, dotted lines, obtained via the fixed LUMA responsiveness ratio, on a log scale, for 80% power, with a two-sided test at the 5% significance level. Marks indicate three scenarios for the treatment effect on MDS-UPDRS Part III: two hypothetical drugs (A, with an effect of 25% reduction — circles and B, with a 50% reduction — triangles), and the effect reported in the specific trials (diamonds), shown for PASADENA and PADOVA; the labelled values (546 and 15,598) are the total sample sizes required to detect these observed effects (33.75% and 15.61% slope reduction, respectively). No treatment-effect information is available for LUMA, and the effect observed in SPARK was negative (−1.64%, Table I) and so falls outside the plotted range — see Methods.

Assuming the same proportional treatment-related slowing on MFC_1_ and the MDS-UPDRS Part III, using MFC_1_ as the endpoint for powering the trial is projected to reduce the required total sample size by approximately 84%. This projection applies the LUMA-derived responsiveness ratio of 2.48 to each of the four reference cohorts under both illustrative treatment-effect scenarios.

Supplementary Fig. S1 presents sample size results including, in addition to the MFC_1_, the MFC_2_ and the Simple Sum v1.2 Score, with calculations performed using the same principles and their responsiveness ratio of 2.61 and 1.43 (Table I), respectively.

**TABLE 1.**
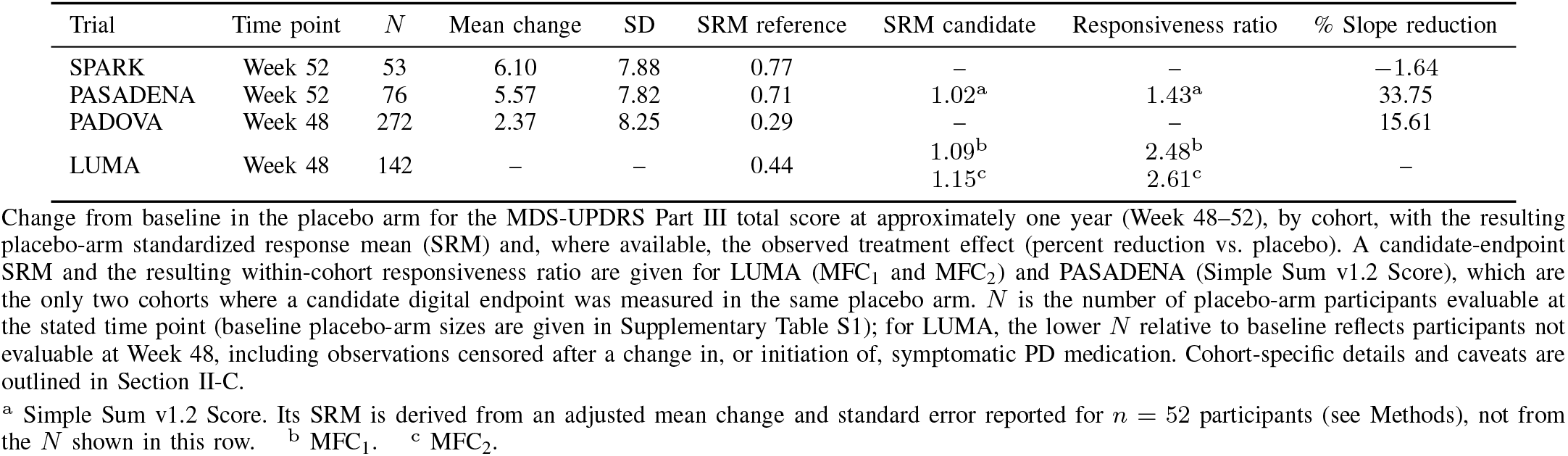
Summary OF THE Needed Estimates FOR THE Four Investigated Trials.

### B. An Interactive Sample Size Calculator using the Responsiveness Ratio approach

The relative-responsiveness approach to the sample size estimation is implemented in an interactive calculator, publicly accessible at https://indiviio.shinyapps.io/SampleSizeCalculator/.

In the calculator, multiple parameters such as the treatment effect and the SRM-based responsiveness ratio can be adjusted, and different data displays can be produced. The calculator is organized around three groups of inputs:

- **Clinical Assumptions** — total enrolled sample size, treatment allocation ratio (from 1:1 up to 4:1), and proportion of enrolled patients expected to be non-evaluable.
- **Endpoint Configuration** — a benchmark, i.e. the reference endpoint in the terminology of the Methods (typically a clinical endpoint), defined by the placebo-arm SRM, and one or more candidate (e.g. digital) endpoints defined by an appropriate responsiveness ratio versus the benchmark (see Methods).
- **Display Settings** — up to three assumed treatment effects, expressed as a % slope reduction, and the sample size range to display.

From these inputs, the calculator simultaneously produces two linked graphical representations for every defined end-point: 1) the statistical power as a function of the assumed % slope reduction, at the specified total sample size; and 2) the statistical power as a function of total sample size, for each of the specified % slope-reduction scenarios.

Both graphical representations are interactive, so hovering over any point of the curve shows the exact power, sample size, and effect-size values.

### C. Illustrative One-Year Trial with 200 Participants

Suppose a clinical development study team is planning a one-year parallel-group trial with a total sample size of 200 early-PD participants with a 1:1 randomization scheme and wants to compare the MDS-UPDRS Part III against the MFC_1_ and MFC_2_ in terms of statistical power. Table I reports the values needed to configure this comparison, all from the LUMA cohort: a benchmark SRM of 0.44 for the MDS-UPDRS Part III, and a responsiveness ratio of 2.48 and 2.61 for the MFC_1_ and MFC_2_ respectively. Entering the benchmark SRM and the Responsiveness Ratios into the Endpoint Configuration panel, together with *N* = 200, 1:1 allocation, and the calculator’s default of 10% for non-evaluable subjects, produces the graphical comparison shown in Fig. 2.

**Fig. 2.**
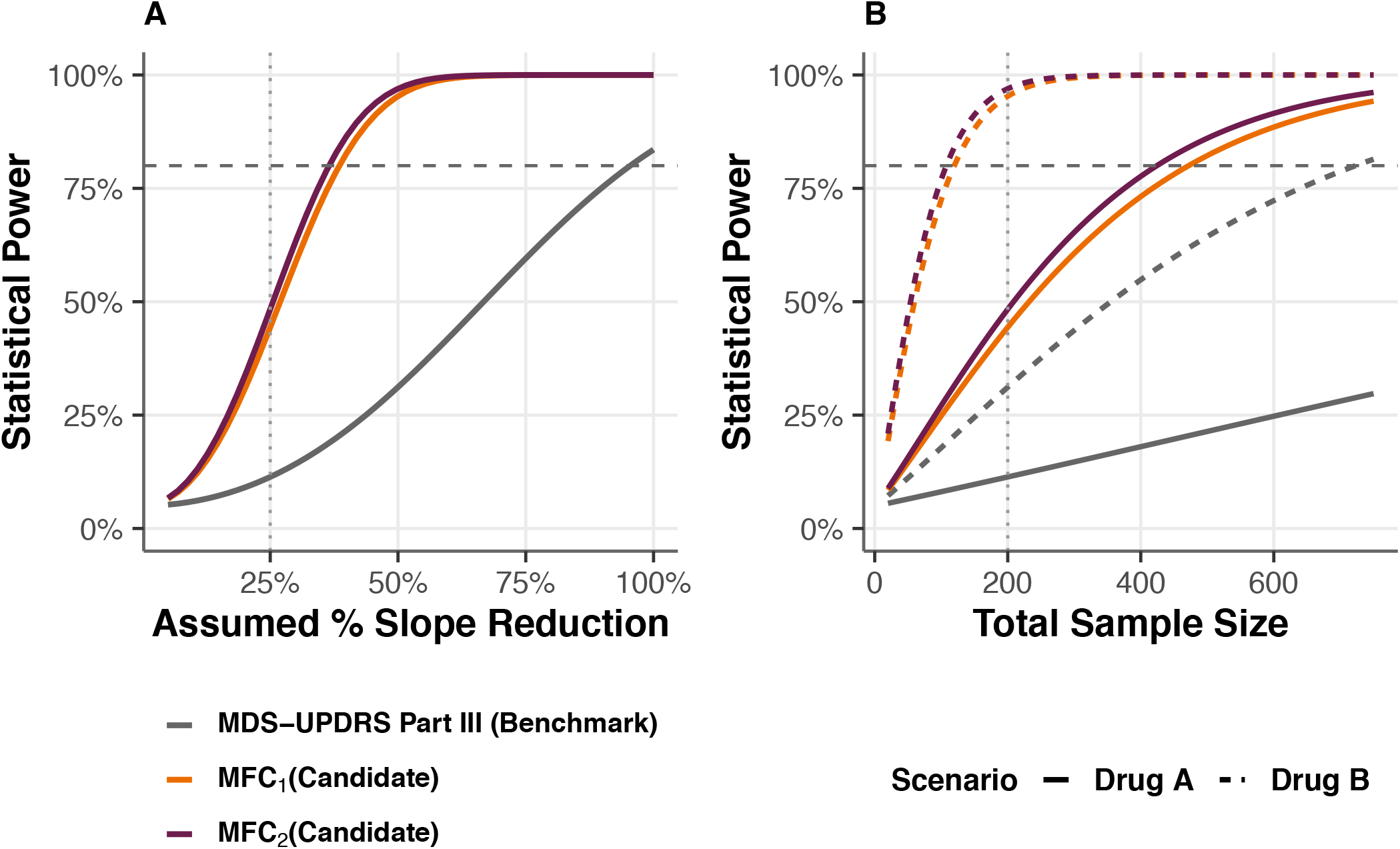
Calculator output for a hypothetical one-year, *N* = 200 trial (1:1 allocation, 10% non-evaluable patients), comparing the MDS-UPDRS Part III to the MFC_1_ and MFC_2_, all anchored on the LUMA placebo arm (Table I). (A) Statistical power vs. assumed % slope reduction on the Benchmark endpoint, at the specified *N* = 200; the vertical dotted line marks the Drug A (25% reduction) scenario. (B) Statistical power vs. total enrolled sample size, for the Drug A (25% reduction) and Drug B (50% reduction) scenarios; the vertical dotted line marks the specified *N* = 200. The horizontal dashed line in both panels marks 80% power.

At *N* = 200, an assumed 50% slowing of progression yields 31% power for the MDS-UPDRS Part III but more than 95% for both MFC_1_ and MFC_2_ (Fig. 2A). Equivalently, reaching the conventional 80% power under the same 50% reduction assumption would require approximately 725 enrolled participants with the MDS-UPDRS Part III as the primary endpoint, versus 120 with the MFC_1_ and 109 with the MFC_2_ (Fig. 2B), consistent with the 2.48-fold and 2.61-fold responsiveness ratios and the sample-size reduction, as already discussed (here additionally accounting for non-evaluable patients). Users can apply the same workflow to any other benchmark/candidate pair and trial design. Changing a benchmark SRM value, a responsiveness ratio, a sample size or an allocation ratio immediately updates both panels.

## IV. DISCUSSION

Data from selected published early PD Phase 2 trials were used to extract responsiveness to disease progression information for the MDS-UPDRS Part III at one year and to estimate the sample size of trials aiming to detect a treatment-related slowing in PD progression.

This work did not aim to present an exhaustive account of all PD trials conducted so far but focused on four recent large Phase 2 trials of candidate disease-modifying therapies for which necessary data were publicly available: SPARK, PASADENA, PADOVA, and LUMA.

Calculations based on placebo-arm data from these Phase 2 studies indicate that trials powered on the MDS-UPDRS Part III would require large samples to detect even a 50% slowing of disease progression. Results point to a strong dependence of the required sample size on the assumed treatment effect, but also on the reference trial used to anchor the assumptions for the MDS-UPDRS Part III, which appear to have a large between-trial variability.

Under the modeled assumptions, MFC_1_ and MFC_2_ offer greater projected statistical power than the MDS-UPDRS Part III for a given target sample size. This projected gain reflects the higher placebo-arm responsiveness to disease progression of MFC_1-2_ endpoints and assumes the same proportional treatment-related slowing on the clinical and digital endpoints.

The superior responsiveness ratios of 2.48 and 2.61 for MFC_1_ and MFC_2_, used for the sample size estimations of trials using these digital outcome measures, are anchored on a single trial (LUMA). Similarly, a superior responsiveness of a digital outcome measure vs. the clinical counterpart was also found for the Simple Sum v1.2 Score derived from PASADENA, with a composite from different assessments and computation rules, which yielded a lower responsiveness ratio of 1.43.

The drivers of the increased responsiveness of MFC_1-2_ are currently being investigated. Because the cross-sectional reliability of the MFCs and of the MDS-UPDRS Part III in early PD is similar [4], [10], the higher administration frequency of the digital measures, with the possibility of aggregating multiple assessments to derive the outcome measures, is likely to play a role in the increase of endpoint responsiveness [11]. For the proposed MFCs in their current form, aggregation over periods of 3 weeks was used.

These projections depend on several assumptions and limitations:

- Stability of the responsiveness in the two treatment arms is assumed. This is likely reasonable for most relevant scenarios, particularly in early PD, because of the rather modest disease progression rates at one year, which implies that the means of the outcome measures at one year are very similar and have a similar variability.
- The change from baseline in the outcome measures is assumed to be linear over time. This is generally a good approximation for at least the relevant time interval (one year). Occasionally, however, some short-term nonlinearities are observed at treatment start. An example is the PADOVA trial [9], where the linear worsening was preceded by a small, short-term improvement: in both arms the mean change stayed at or below baseline for roughly the first 20 weeks. Because the one-year change from baseline then understates the underlying progression rate, the PADOVA-anchored sample sizes reported here are conservative, and such effects would require a slight adaptation of the sample size formulae.
- Sensitivity to change does not necessarily translate to sensitivity to treatment effects, and the treatment effect on slope reduction may be different for different measures. No data on the treatment effect observed in LUMA are publicly available, beyond the communication that the trial was negative; data on the sensitivity of the MFCs to treatment effects are therefore lacking. In the development of the Motor Functional Composite, care was taken to have a broad representation of relevant motor function domains (with an emphasis on gait), so that it is reasonable to expect treatment effects (slope reduction) to be in a similar range for the two measures, although the presence of potential differences remains to be formally investigated.
- Because the novel digital endpoint is based on a single-trial analysis, information is lacking about the between-trial variability: even if one is able to identify suitable references for the design elements of a new trial (e.g., selection criteria, duration, geographies, mechanism of action of the drug), the generalizability of the predictions of outcomes to future trials remains to be demonstrated.
- Only one type of endpoint (mean change from baseline at one year) was considered. For early phase studies, a primary analysis after one year of treatment reflects a typical scenario. However, Phase 3 studies are typically longer and may use other types of endpoints (such as time to the first confirmed progression event), but data on long-term responsiveness, or other endpoints, remain to be explored.

Taken together, these results pave the way for a potential shift in how early-phase Parkinson’s disease trials are designed and powered to evaluate the proof-of-concept of disease modifying interventions. In the illustrative one-year trial with 200 participants, equal treatment allocation, and 10% non-evaluable participants, an assumed 50% slowing of progression yields projected statistical power of 31% with the MDS-UPDRS Part III and more than 95% with either MFC_1_ or MFC_2_. Alternatively, achieving 80% power under the same assumptions would require approximately 725 enrolled participants with the MDS-UPDRS Part III, compared with 120 with MFC_1_ or 109 with MFC_2_. If adopted at scale, this notable gain in statistical efficiency could let sponsors test more candidate therapies in smaller trials, while reaching end-of-phase 2 go-no-go decisions with greater confidence. If confirmed across additional trials and therapies, such MFC endpoints have the potential to meaningfully improve the PD trial ecosystem, enabling adequately powered proof-of-concept studies with fewer participants and supporting more efficient evaluation of candidate disease-modifying treatments.

## Data Availability

All data used in this study are publicly available aggregate summary statistics from the cited sources: SPARK (NCT03318523; Lang et al., N Engl J Med 2022, https://doi.org/10.1056/NEJMoa2203395, and ClinicalTrials.gov results disclosure), PASADENA (NCT03100149; Pagano et al., N Engl J Med 2022, https://doi.org/10.1056/NEJMoa2202867, and ClinicalTrials.gov results disclosure; Taylor et al., npj Digit Med 2025, https://www.nature.com/articles/s41746-025-01572-8), PADOVA (NCT04777331; Nikolcheva et al., Lancet 2026) and LUMA (NCT05348785; Llorens Arenas et al., International Congress of Parkinson's Disease and Movement Disorders 2026, poster 742). All values extracted from these sources are contained in the manuscript (Table I and Supplementary Table S1). The sample size calculations are implemented in a publicly accessible interactive application at https://indiviio.shinyapps.io/SampleSizeCalculator/.

## Declarations

### Ethics Statement

This study is a secondary analysis of publicly available, aggregate summary statistics reported in peer-reviewed publications, trial results disclosures and conference presentations. No individual participant data were accessed, and no new data were collected; ethical approval and informed consent were therefore not required for this analysis. Each source trial was conducted under its own ethics approvals and informed-consent procedures, as described in the original publications.

### Clinical Trial Registration

This study is not a clinical trial and was not prospectively registered. It uses published summary data from the following registered trials: SPARK (NCT03318523), PASADENA (NCT03100149), PADOVA (NCT04777331) and LUMA (NCT05348785).

### Data Availability

All data used in this study are publicly available in the cited sources [4], [5], [7]–[9]; the values extracted from each source are reported in Table I and Supplementary Table S1.

## Supplementary Materials

### Baseline Characteristics of Placebo Arms

To contextualize the progression rates in the different cohorts, Supplementary Table S1 summarizes the baseline *Sample-Size Reduction from the Smartphone-Based Motor Functional Composite (MFC*_*1-2*_*) and the Simple Sum v1*.*2 Score*

**TABLE S1.** Baseline Characteristics OF THE Placebo Arm BY Trial.

| Trial | *N* | Age, years, mean (SD) | MDS-UPDRS III baseline (OFF), mean (SD) | Time since diagnosis, months, mean (SD) | Pre-treated with symptomatic medication, % | Hoehn and Yahr, % by stage |
| --- | --- | --- | --- | --- | --- | --- |
| SPARK | 100 | 61.0 (8.4) | 22.2 (9.3) | 8.4 (7.2) | ~20<sup>a</sup> | Stage 1: 29%; Stage 1.5: 3%; Stage 2: 64%; Stage 2.5: 3%; Stage 3: 1% |
| PASADENA | 105 | 59.9 (8.7) | 21.5 (9.1) | 10.0 (6.8) | 36.2<sup>b</sup> | Stage 1: 19%; Stage 2: 81% |
| PADOVA | 293 | 64.4 (7.5) | 24.6 (10.8) | 18.4 (9.0) | 100 | Stage 1: 14%; Stage 2: 85%; Stage 3: 1% |
| LUMA<sup>c</sup> | 303 | 60.7 (9.6) | 19.2 (7.8) | N.A.<sup>d</sup> | N.A.<sup>d</sup> | N.A.<sup>d</sup> |

<sup>a</sup> Approximately 20% of the overall enrolled SPARK population reported having taken some PD medication before study entry (i.e. ~80% treatment-naïve); this figure was reported for the full trial, not broken out for the placebo arm specifically. They were requested to stop the PD symptomatic medication 30 days before the first dose.
<sup>b</sup> Monoamine oxidase B inhibitors, the only symptomatic treatment permitted before entry in PASADENA.
<sup>c</sup> As of 4th of October 2026, the values for LUMA were not reported for the whole trial but only for those who also took part in digital assessments and are also evaluable for the MFC; it is assumed that the overall placebo arm has similar values.
<sup>d</sup> As of 4th of October 2026, data were not reported. Inclusion criteria: “Were diagnosed with Parkinson’s disease within the last 2 years and were at least 30 years old when you were diagnosed”.
*N* is the number of randomized placebo-arm participants at baseline; the number evaluable at the one-year time point is given in Table I.

The Simple Sum v1.2 Score, the smartphone-derived composite measured alongside the MDS-UPDRS Part III in the PASADENA placebo arm, can be translated into a projected sample-size curve in exactly the same way as the MFC_1_ and MFC_2_, by substituting its own responsiveness ratio (1.43, PASADENA-anchored) for the LUMA-anchored ratio (2.48 and 2.61) used for the Motor Functional Composite (MFC_1_ and MFC_2_).

Estimating the SRM for the Simple Sum v1.2 Score required the extraction of information from a figure in Taylor et al. [5]. While the mean change from baseline (4.4) could be read off reliably, the standard error at that specific point was difficult to pin down precisely; two independent AI-assisted readings of the same figure (Claude Sonnet 5 and Gemini 3.6) gave estimates of 0.5 and 0.7, respectively. In the absence of a principled way to choose between them, the midpoint, 0.6, was used as the primary estimate throughout this paper. Propagating the two extremes through the same SRM and responsiveness-ratio calculation gives an SRM of 1.22 for SE = 0.5 and 0.87 for SE = 0.7, corresponding to responsiveness ratios of 1.71 and 1.22 respectively. The adopted value of 1.43 (Table I) is the ratio implied by the midpoint SE; because the ratio scales as 1*/SE*, it is not the midpoint of the 1.22– 1.71 range. This range of uncertainty is illustrated as error bars on the Simple Sum v1.2 Score bars in Supplementary Fig. S1, translating directly into a total-sample-size range at the 25% slope-reduction scenario, in contrast to the single demographic and clinical characteristics of the placebo control arm in the respective studies. point estimates available for the other endpoints.

Supplementary Fig. S1 illustrates how the responsiveness ratios translate into projected sample size requirements across the four trial cohorts, assuming a 25% slowing of disease progression and 80% statistical power. Under these assumptions, using MFC_1-2_ in place of the MDS-UPDRS Part III is projected to reduce the required total sample size by approximately 84–85%. Using the Simple Sum v1.2 Score is projected to reduce the required total sample size by approximately 51%. These three digital endpoints therefore offer potentially substantial gains in statistical efficiency, with the larger projected reductions for MFC_1-2_ reflecting higher placebo-arm responsiveness ratios relative to the MDS-UPDRS Part III (2.48–2.61 for MFC_1-2_ versus 1.43 for the Simple Sum v1.2 Score; Table I). For each endpoint, the proportional reduction is consistent across cohorts because a fixed responsiveness ratio is applied, whereas the absolute sample size requirements vary with each cohort’s underlying MDS-UPDRS Part III responsiveness.

This comparison inherits the caveats already noted for the Simple Sum v1.2 Score SRM: it was read off a published figure (see above), and rests on a single external cohort without confirmation within the same trial in a held-out dataset (as done for the MFC_1-2_). The Simple Sum v1.2 Score values in Supplementary Fig. S1 should therefore be read as a conservative and currently less certain estimate of the achievable sample-size reduction than the MFC_1-2_ values, not as a like-for-like comparison to it.

**Fig. S1.**
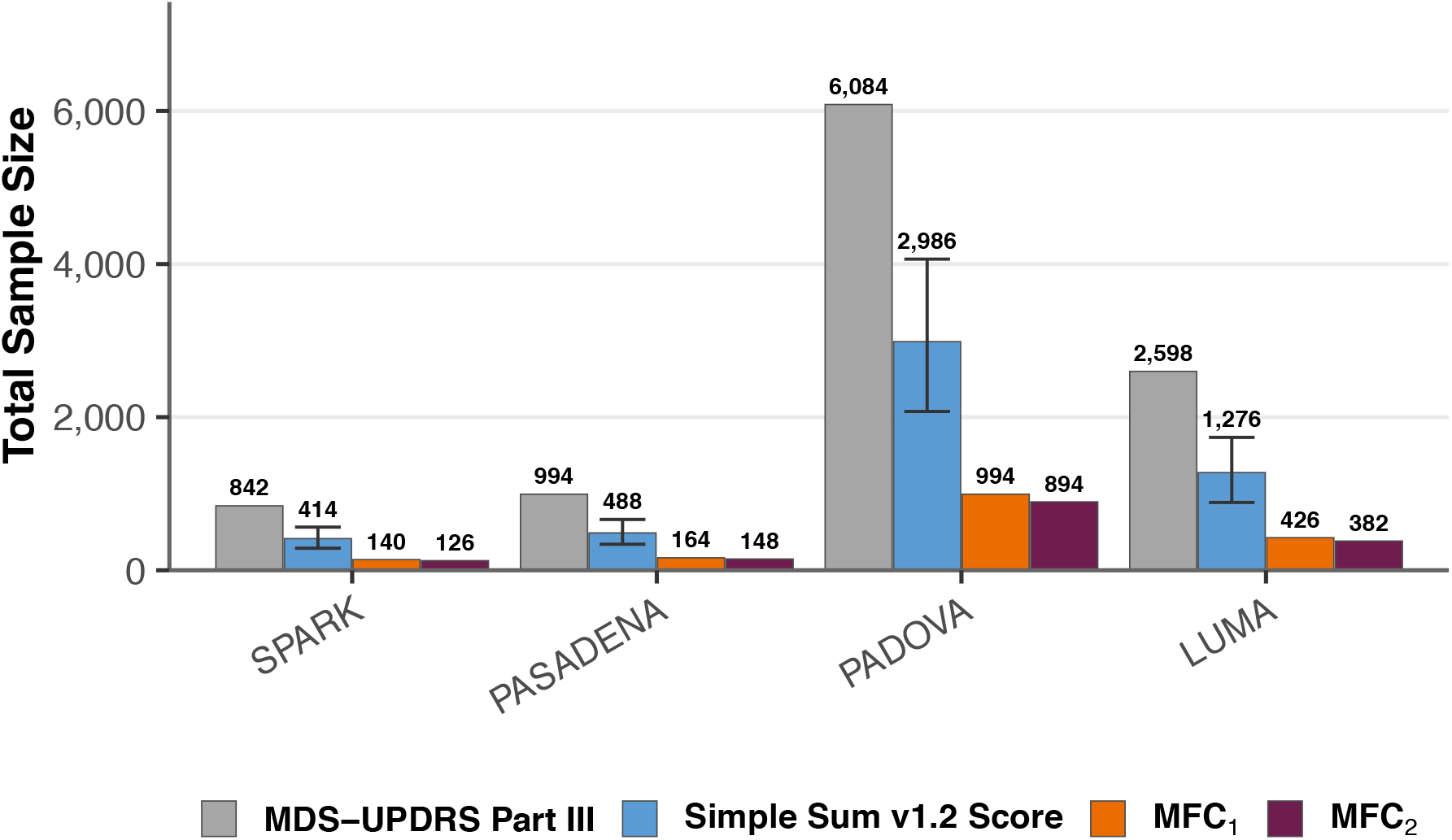
Total sample size (80% power, two-sided test, *α* = 0.05) required using the MDS-UPDRS Part III (reference), the Simple Sum v1.2 Score, and the Motor Functional Composite (MFC_1_ and MFC_2_), for the 25% assumed slope-reduction scenario, by trial cohort. The Simple Sum v1.2 Score, MFC_1_ and MFC_2_ bars are each obtained by applying a fixed, endpoint-specific responsiveness ratio (PASADENA for the Simple Sum v1.2 Score, LUMA for the MFC_1-2_) to every cohort’s reference-endpoint effect size. Error bars on the Simple Sum v1.2 Score reflect the uncertainty in its figure-derived SRM (see text).

